# NHS staff: Sickness absence and intention to leave the profession

**DOI:** 10.1101/2024.08.05.24311412

**Authors:** Lauren J Scott, Danielle Lamb, Chris Penfold, Maria Theresa Redaniel, Nora Trompeter, Paul Moran, Rupa Bhundia, Neil Greenberg, Rosalind Raine, Simon Wessely, Ira Madan, Peter Aitken, Anne Marie Rafferty, Sarah Dorrington, Richard Morriss, Dominic Murphy, Sharon A.M. Stevelink

**Affiliations:** National Institute for Health Research Applied Research Collaboration West (NIHR ARC West), University Hospitals Bristol and Weston NHS Foundation Trust, Bristol, UK; Population health Sciences, Bristol Medical School, University of Bristol, Bristol, UK; Department of Applied Health Research, University College London, London, UK; Institute of Psychiatry, Psychology & Neuroscience, King’s College London, London, UK; Guy’s and St Thomas’ NHS Trust, London, UK; Sussex Partnership NHS Foundation trust, Sussex, UK; Florence Nightingale Faculty of Nursing, Midwifery & Palliative Care, King’s College London, London, UK; University of Nottingham, Nottingham, UK

**Keywords:** NHS, Workforce, Staffing, Predictors, Exposures, Sickness absence, intention to leave

## Abstract

**Objective:** To determine key workforce variables (demographic, health and occupational) that predicted NHS staff’s 1) absence due to illness (both COVID-19 and non-COVID-19 related) and 2) expressed intention to leave their current profession.

**Methods:** Staff from 18 NHS Trusts were surveyed between April 2020 and January 2021, and again approximately 12months later. Logistic and linear regression were used to explore relationships between baseline exposures and 12-month outcomes.

**Results:** We included 10,831 participants for analysis. At 12-months, 20% stated they agreed or strongly agreed they were actively seeking employment outside their current profession; 24% said they thought about leaving their profession at least several times per week. Twenty-percent of participants took 5+ days of work absence due to non-COVID-19 sickness in the 12-months between baseline and 12-month questionnaire; 14% took 5+ days of COVID-19 related sickness absence. Sickness absence (COVID-19 and non-COVID-19 related) and intention to leave the profession (actively seeking another role and thinking about leaving) were all more common among NHS staff who were younger, in a COVID-19 risk group, had a probable mental health disorder, and who did not feel supported by colleagues and managers.

**Conclusions:** There were several factors which affect both workforce retention and sickness absence. Of particular interest because they are modifiable, are the impact of colleague and manager support. The NHS workforce is likely to benefit from training managers to speak with and support staff, especially those experiencing mental health difficulties. Further, staff should be given sufficient opportunities to form and foster social connections.

**What is already known on this topic:**

- The ability of the NHS to provide a good service in a timely manner is under more pressure and strain than ever before, and therefore the retention and health of current staff is incredibly important.

**What this study adds:**

- We included survey data collected on 10,831 NHS staff across 18 Trusts in England between April 2020 and February 2022.
- Sickness absence and intention to leave the profession were more common among NHS staff who were younger, in a COVID-19 risk group, had a probable mental health disorder, and who did not feel supported by colleagues and managers.

**How this study might affect research, practice or policy:**

- This study suggests that in order to improve staff retention and reduce staff sickness, the NHS workforce is likely to benefit from training managers to speak with and support staff, especially those experiencing mental health difficulties.
- Further, staff should be given sufficient opportunities to form and foster social connections and reflect on the challenges of the work they do together.

## Introduction

The National Health Service (NHS; a government funded universal healthcare provider in the UK), and in particular its staffing, is a regular topic of UK media and political debate. The reporting of this topic frequently states that the ability of the NHS to provide a good service in a timely manner is under more pressure and strain than ever before (1).

There are many potential reasons for the current workforce difficulties faced by the NHS. These include the disruption caused by the COVID-19 pandemic and ongoing economic difficulties facing the UK (2). Sickness absence rates are reported to be at an all-time high (3, 4), as are the number of staff intending to leave or leaving the NHS (5). Poor staffing levels mean remaining staff have more work to do, with 71% of NHS staff surveyed stating that they do not have as much time with patients as they would like (6). This can be a source of workplace stress which can become sickness absence; the Health and Safety Executive reports that, across all sectors, mental ill-health is the most common reason for sickness absence from work (7, 8). The reduction of sickness and absence is a key ambition of the long term NHS Workforce plan (9).

The NHS CHECK study was developed early in the pandemic to try to understand the mental health and well-being of clinical and non-clinical NHS healthcare workers across 18 Trusts in England (10). Data were collected early in the pandemic, and again 6 and 12 months after the baseline questionnaire (10). While the original focus of these questionnaires was on the mental health implications of the pandemic for NHS staff, the data provides an opportunity to assess a wider range of aspects regarding the wellbeing of staff working in the NHS.

The aim of this study was to determine key baseline workforce variables (demographic, health and occupational) that predicted 1) NHS staff absence due to illness (both COVID-19 and non-COVID-19 related) and 2) the expressed intention to leave their current profession, 12-months later.

## Methods

### Setting and participants

Eighteen English NHS Trusts were selected for this study, chosen for diversity in geographical location, urban and rural settings, and acute and mental health Trusts (10). All Trust staff were eligible to participate, including clinical (e.g. doctors, nurses, etc) and non-clinical (e.g. administration roles, human resources roles, porters and cleaners) staff, and staff on any contract type. Trusts were invited to participate via direct emails to senior leadership teams, building upon the research team’s professional network.

### Data collection

Once recruited, Trusts invited all staff to participate in an online survey. The study was promoted via several routes, and both individuals and Trusts were incentivised with prizes (10). The baseline data collection period was April 2020 to January 2021. Follow-up data were collected approximately 6 and 12 months after each individual completed the baseline survey. Twelve-month data collection was completed between May 2021 and February 2022 (10).

This paper focuses on baseline exposures and 12-month outcomes, and only includes participants who completed, at least in part, the baseline and 12-month questionnaires. All questionnaires were split into two parts; once a participant had completed the first part (“short questionnaire”), they were asked if they wished to complete further questions in the second part (“long questionnaire”). This split was to encourage staff to participate even if they were short of time; 54% of participants who completed the short 12-month questionnaire also completed the long questionnaire. The intention to leave outcomes, and the COVID-19 risk group exposure variable (see below), were recorded in the long questionnaires.

### Outcomes

The four outcomes were responses to the following statements and questions, recorded in the 12-month questionnaire:

- “I am actively seeking employment outside my current profession or occupation” (Strongly disagree, disagree, neither agree or disagree, agree, strongly agree). Dichotomised as: agree/strongly agree vs. the other three categories.
- “How often do you think about leaving your current profession or occupation?” (never, several times a year, several times a month, several times a week, everyday). Dichotomised as: several times a month/week/daily vs. the other two categories.
- “How many days in total in the last 12 months have you been absent from work due to ill health that is not related to COVID-19 (it is fine to estimate)?”
- “How many days in total have you been absent from work due to COVID-19 (it’s fine to estimate)?”

### Exposure variables

The exposure variables of interest were recorded in the baseline questionnaire and were selected *a priori* based on expert opinion:

- Age (categorised as ≤30, 31-40, 41-50, 51-60, and ≥61 years)
- Gender (male, female, other, prefer not to say)
- Ethnicity (white, black/African/Caribbean, Asian, mixed/multiple, other)
- Clinical role (doctor, nurse/midwife, other clinical, non-clinical),
- COVID-19 risk group membership (yes, no; most yeses were for pre-existing health conditions, but also included the elderly, pregnant or other risk reasons)
- Mental health status according to the general health questionnaire-12 (11) (GHQ12; no common mental health disorder [GHQ12<4], probable common mental health disorder [GHQ12≥4])
- Type of Trust (acute or mental health)
- Redeployed outside usual role (yes, no)
- Felt supported by colleagues (not at all, a little bit, moderately, quite a bit, extremely)
- Felt supported by manager (not at all, a little bit, moderately, quite a bit, extremely).

In addition, a covariable for season in which the 12-month questionnaire was completed was included in all models, to account for seasonal differences in hospital admissions and therefore staff workload (spring/summer [May-Aug 2021], autumn [Sep-Nov 2021], winter [Dec 2021-Feb 2022].

For the GHQ12, the robustness of the measure has been established even where response options differ slightly from the original, as was the case in this study where there was a small typographical error in one response option of one GHQ-12 item (12). If a participant had completed 10 or 11 out of the 12 questions, then the most common score across the cohort for the given question was imputed for the remaining one or two questions. The overall score was calculated in the standard way.(11, 13) This overall score (min=0, max=12) was dichotomised according to standard guidelines as <4 (no mental health disorder) and ≥4 (probably common mental health disorder).

### Statistical analysis

Continuous data are summarised using means and standard deviations (SDs), or medians and interquartile ranges (IQRs) if distributions are highly skewed. Categorical data are summarised as numbers and percentages.

Predictors of intention to leave current profession were explored using logistic regression. We undertook two sets of statistical models: one for actively seeking employment outside their current profession and one for thinking about leaving their current profession. Each exposure variable was fitted in the models in turn, adjusting for age, gender ethnicity, and season of 12-month questionnaire completion, as fixed effects. Odds ratios (ORs), 95% confidence intervals (CIs) and p-values are presented.

Due to the large number of participants with no sick days, predictors of the number of non-COVID-19 related and COVID-19 related sick days were explored using two-part modelling. For each of the outcomes, logistic regression models were fitted with the outcome of any vs. no sick days and linear regression models were fitted only for the participants who had at least one sick day. These were fitted using the ‘twopm’ command in Stata, with the ‘suest’ option to combine the estimation results of the two parts of the model to derive a simultaneous variance matrix. Models were fitted for each exposure variable adjusting for demographic variables and season, as above. For the logistic regression parts, ORs, 95% CIs and p-values are presented. Model fit of linear models were explored graphically, and the continuous part of all outcome variables were log transformed for best fit; estimates are presented as geometric mean ratios (GMRs), 95% CIs and p-values.

For categorical variables with more than two categories, p-values for the overall effect of the variable are also presented. Due to small numbers, mixed and other ethnicity categories were combined, and participants whose gender was other or prefer not to say were excluded in all models.

Analyses are all based on complete case data. To explore the effects of missing data, we present baseline characteristics for the full baseline cohort, as well as those who completed the short and long parts of the 12-month questionnaire in Table 1, and present those with complete outcome data for each of the four outcomes in Supplementary Table S1. Further, we explored which exposure variables predicted missingness at 12-months, taking the whole baseline cohort and predicting missingness in each of the four specific outcomes. These were modelled using logistic regression, adjusted the same as the outcome models. Findings from these analyses are presented in supplementary Table S2 and discussed in the limitations section in the discussion.

**Table 1:**
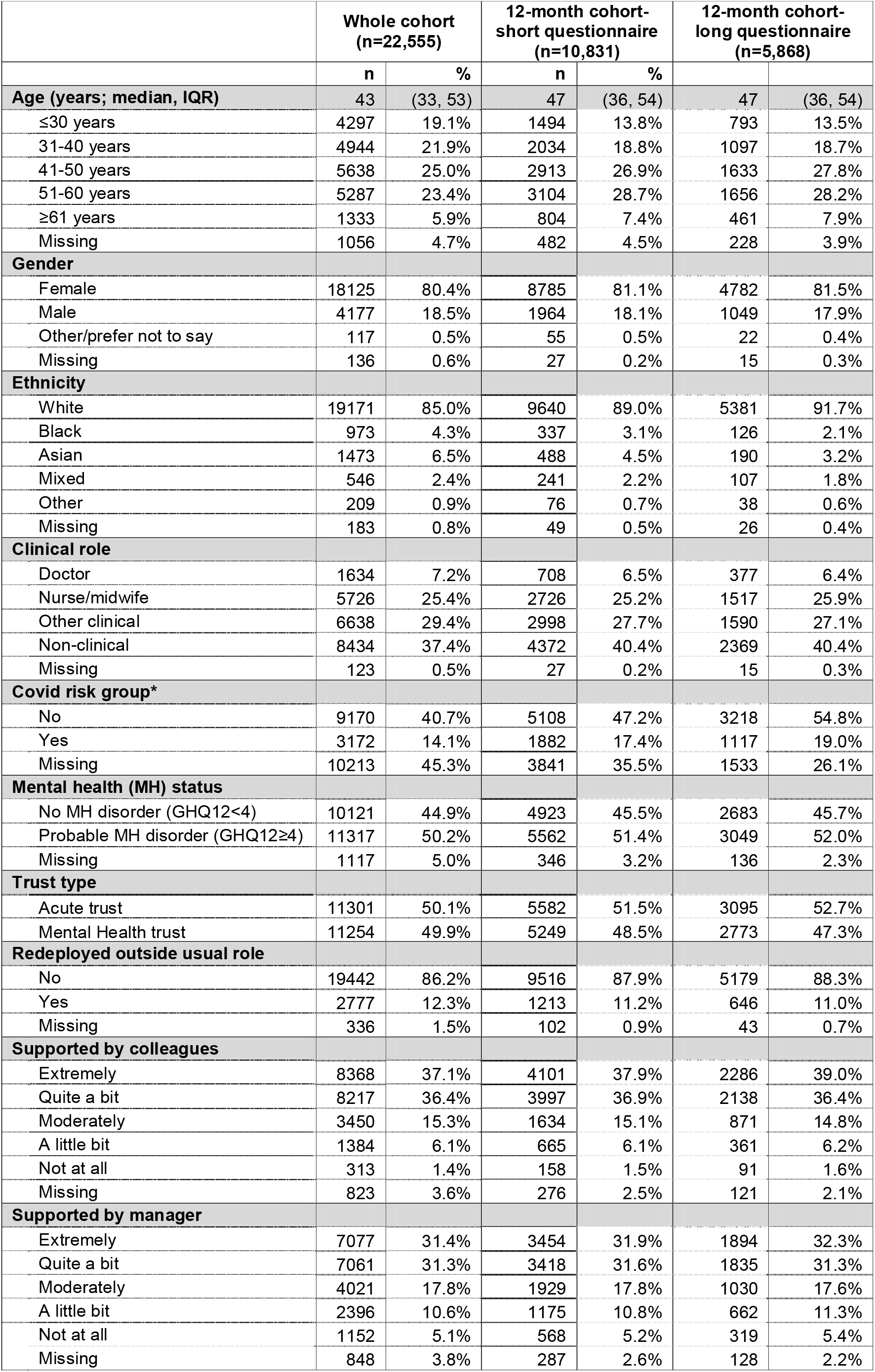
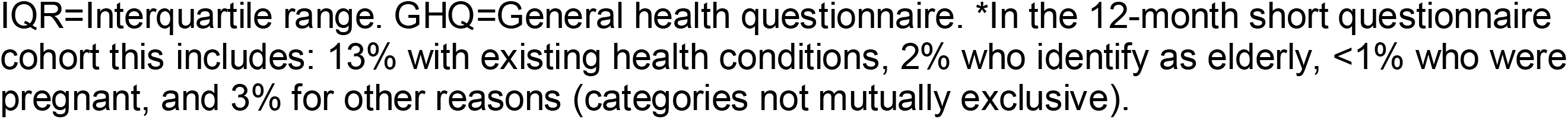
Participant baseline characteristics.

Ethical approval for the NHS CHECK study was granted by the Health Research Authority (reference: 20/HRA/210, IRAS: 282686) and local Trust Research and Development. All data management and analyses were performed in Stata version 17.0.

## Results

### Baseline data

Valid responses to the baseline questionnaire (collected April 2020-January 2021) were received from 22,555 participants, a 15% response rate from a potential denominator of 152,268 employees. After excluding participants who did not complete the 12-month questionnaire, we included 10,831 participants for analysis (of these, 5,868 completed the long questionnaire and therefore had the opportunity to complete the intention to leave questions). The median age of participants was 47 years (IQR 36 to 54), 81% were female, and 89% were white. 60% of participants held clinical roles, comprising 7% doctors, 25% nurses/midwives and 28% other clinical roles; 40% held a non-clinical role. Demographics were similar between those who completed the baseline questionnaire, and the short and long 12-month questionnaires (Table 1).

### 12-month data

8% of participants (1,169/5,868) stated they strongly agreed that they were actively seeking employment outside their current profession; 12% (710/5,868) agreed, 18% (1,052/5,868) neither agreed or disagreed, 18% (1,067/5,868) disagreed, 43% (2,528/5,868) strongly disagreed and 1% (52/5,868) did not answer this question. 12% of participants (731/5,868) said they thought about leaving their profession every day, 12% (726/5,868) thought about it several times per week, 18% (1,044/5,868) several times per month, 29% (1,694/5,868) several times per year, 28% (1,631/5,868) never thought about it, and 1% (42/5,868) did not answer this question.

39% of participants (4,221/10,831) took no absence from work due to non-COVID-19 illness in the 12-month period before completing the questionnaire, 18% (2,001/10,831) took 1-4 days, 7% (746/10,831) 5-9 days, 3% (349/10,831) 10-14 days, 10% (1,041/10,831) 15+ days, and 23% (2,473/10,831) did not answer this question. 61% of participants (6,581/10,831) had no work absence due to COVID-19, 2% (170/10,831) took 1-4 days of COVID-19 related absence, 3% (339/10,831) 5-9 days, 5% (556/10,831) 10-14 days, 5% (558/10,831) 15+ days, and 24% (2,597/10,831) did not answer this question.

### Actively seeking employment outside current profession

Factors associated with participants reporting actively seeking employment outside their current profession were: male gender (OR=1.18, 95%CI 1.00-1.40), being in a COVID-19 risk group (OR=1.34, 95%CI 1.12-1.59), having a probable mental health disorder (OR=1.89, 95%CI 1.64-2.18), and having less colleague (p<0.001) and manager (p<0.001) support (Table 2). Older participants were less likely to be seeking employment outside their current profession (p=0.023). Clinical role was associated (p<0.001), such that doctors were less likely than non-clinical participants to be seeking employment outside their current profession (OR=0.38, 95%CI 0.27-0.55; Table 2).

**Table 2:** Actively seeking a job outside my current profession or occupation (strongly agree/ agree vs. neither agree or disagree/ disagree/ strongly disagree)

|  | Number with outcome |  | Adjusted analysis model |  |  |
| --- | --- | --- | --- | --- | --- |
| Baseline predictor | n | % | Odds ratio | 95% CI | P-value |
| Age: |  |  |  |  | 0.023 |
| ≤30 years | 177 | 22.5% | 1 |  |  |
| 31-40 years | 237 | 21.8% | 0.96 | 0.77 to 1.20 | 0.721 |
| 41-50 years | 326 | 20.2% | 0.87 | 0.71 to 1.07 | 0.180 |
| 51-60 years | 300 | 18.4% | 0.78 | 0.63 to 0.97 | 0.022 |
| 61+ years | 73 | 16.3% | 0.67 | 0.49 to 0.90 | 0.009 |
| Gender: |  |  |  |  |  |
| Female | 889 | 19.5% | 1 |  |  |
| Male | 224 | 22.2% | 1.18 | 1.00 to 1.40 | 0.048 |
| Ethnicity: |  |  |  |  | 0.097 |
| White | 1005 | 19.6% | 1 |  |  |
| Black/African/Caribbean | 31 | 26.3% | 1.45 | 0.95 to 2.20 | 0.082 |
| Asian | 49 | 26.5% | 1.38 | 0.98 to 1.93 | 0.062 |
| Mixed/multiple/other | 28 | 20.1% | 0.98 | 0.64 to 1.50 | 0.933 |
| Clinical role: |  |  |  |  | <0.001 |
| Non-clinical | 470 | 21.1% | 1 |  |  |
| Doctor | 37 | 10.2% | 0.38 | 0.27 to 0.55 | <0.001 |
| Nurse/midwife | 314 | 21.6% | 1.03 | 0.87 to 1.21 | 0.750 |
| Other clinical | 290 | 19.1% | 0.85 | 0.72 to 1.01 | 0.064 |
| COVID risk group: |  |  |  |  |  |
| No | 607 | 19.8% | 1 |  |  |
| Yes | 253 | 23.9% | 1.34 | 1.12 to 1.59 | 0.001 |
| Mental health (MH) status: |  |  |  |  |  |
| No MH disorder (GHQ12<4) | 374 | 14.7% | 1 |  |  |
| Probable MH disorder (GHQ12≥4) | 718 | 24.6% | 1.89 | 1.64 to 2.18 | <0.001 |
| Type of trust staff work for: |  |  |  |  |  |
| Acute | 593 | 20.3% | 1 |  |  |

| Baseline predictor | Number with outcome |  | Adjusted analysis model |  |  |
| --- | --- | --- | --- | --- | --- |
|  | n | % | Odds ratio | 95% CI | P-value |
| Mental health | 520 | 19.7% | 0.97 | 0.85 to 1.12 | 0.687 |
| Redeployed outside usual role: |  |  |  |  |  |
| No | 964 | 19.6% | 1 |  |  |
| Yes | 144 | 23.2% | 1.21 | 0.99 to 1.48 | 0.061 |
| Felt supported by colleagues: |  |  |  |  | <0.001 |
| Extremely | 331 | 15.3% | 1 |  |  |
| Quite a lot | 413 | 20.1% | 1.37 | 1.17 to 1.61 | <0.001 |
| Moderately | 204 | 24.5% | 1.77 | 1.45 to 2.15 | <0.001 |
| A little | 117 | 34.2% | 2.86 | 2.22 to 3.68 | <0.001 |
| Not at all | 31 | 37.3% | 3.19 | 2.01 to 5.07 | <0.001 |
| Felt supported by manager: |  |  |  |  | <0.001 |
| Extremely | 254 | 14.2% | 1 |  |  |
| Quite a lot | 306 | 17.4% | 1.26 | 1.05 to 1.51 | 0.014 |
| Moderately | 226 | 23.1% | 1.80 | 1.47 to 2.20 | <0.001 |
| A little | 198 | 31.3% | 2.76 | 2.23 to 3.42 | <0.001 |
| Not at all | 112 | 37.1% | 3.58 | 2.73 to 4.69 | <0.001 |

CI=Confidence interval. GHQ12=General health questionnaire. All analyses are adjusted for age, gender, and ethnicity. Numbers and analyses only include patients with complete baseline age, gender and ethnicity data, complete outcome data, and who completed the long questionnaire.

### Regularly thinking about leaving current profession

Clinical role (p<0.001; in particular being a nurse/midwife compared to a non-clinical role [OR=1.54, 95%CI 1.35-1.75]), being in a COVID-19 risk group (OR=1.19, 95%CI 1.02-1.37), having a probable mental health disorder (OR=2.28, 95%CI 2.04-2.55), and less colleague (p<0.001) and manager support (p<0.001) increased the odds of regularly thinking about leaving their current profession (Table 3). Older participants were less likely to regularly think about leaving their role than younger participants (p=0.001; Table 3).

**Table 3:**
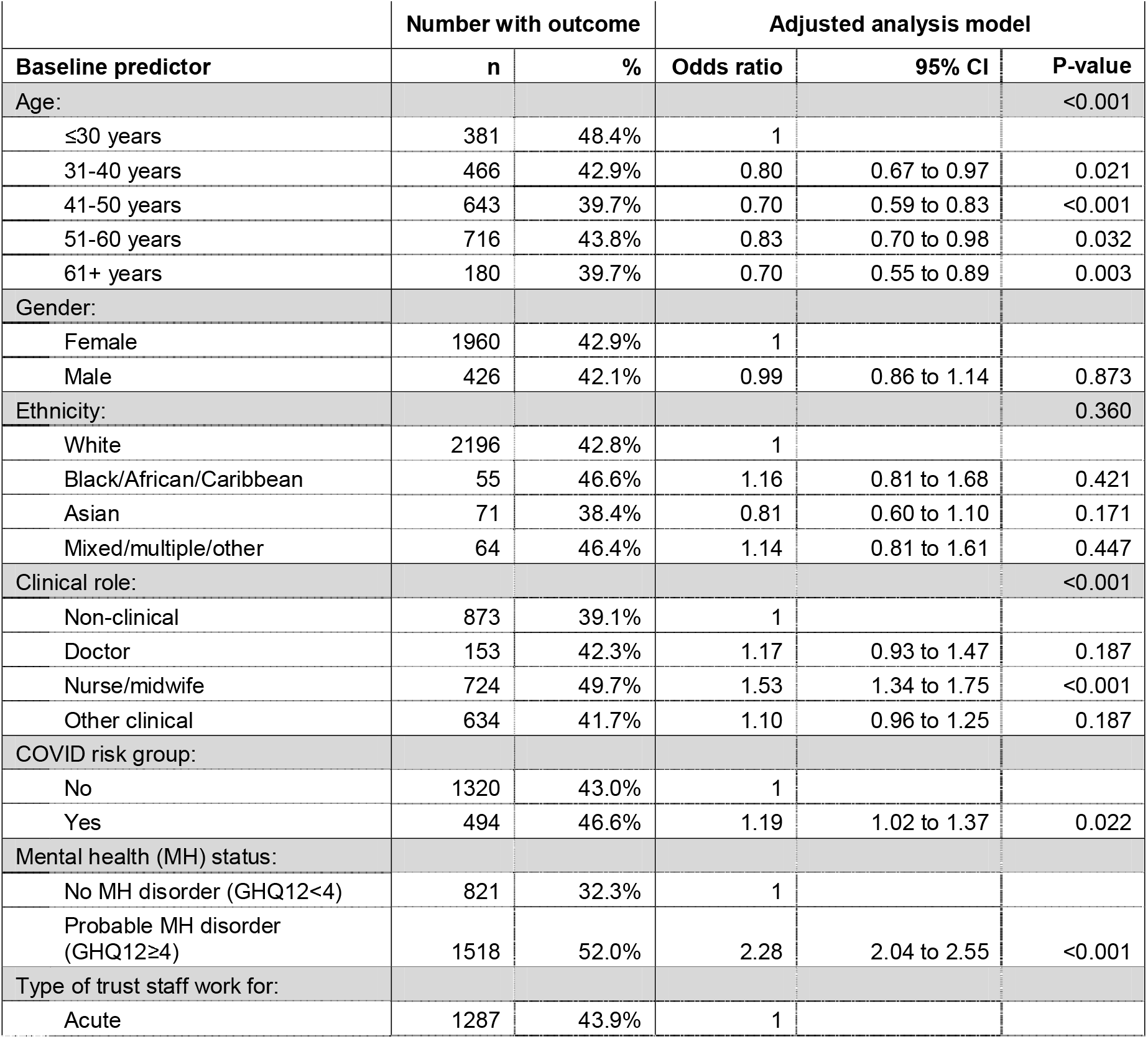
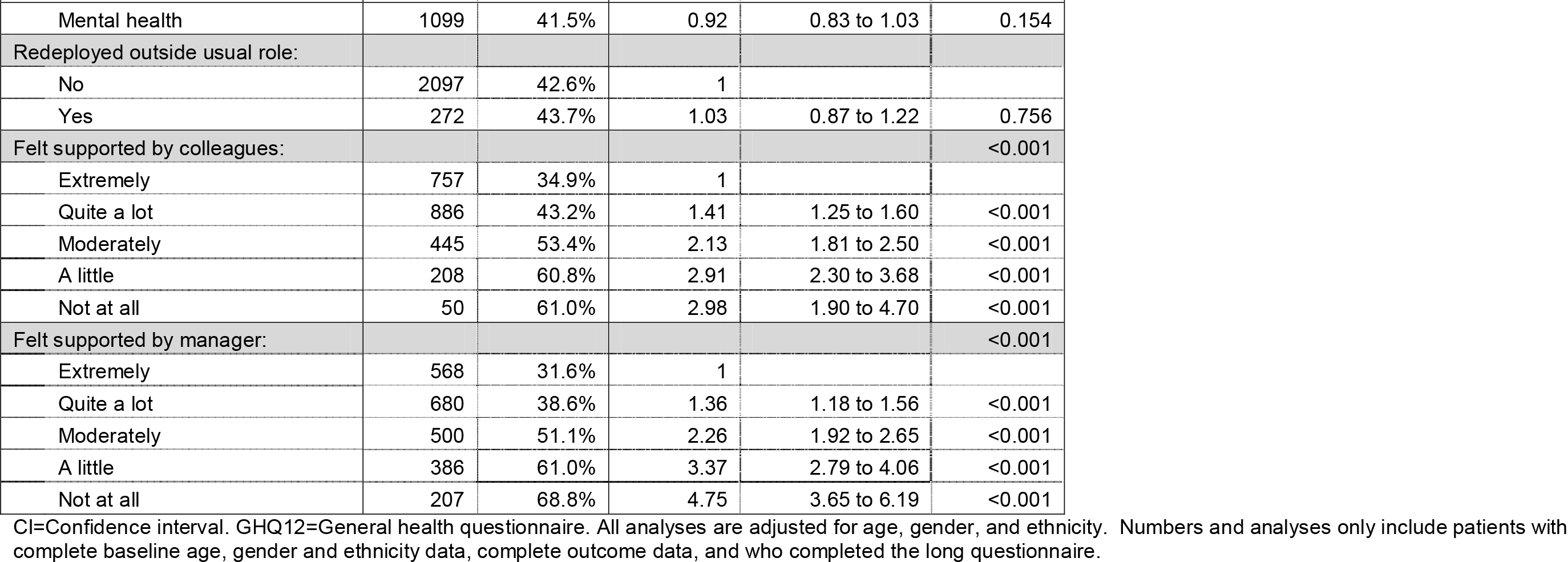
Thinking about leaving your current profession or occupation (Every day/ several times a week/ several times a month vs. several times a year/ never)

|  | Number with outcome |  | Adjusted analysis model |  |  |
| --- | --- | --- | --- | --- | --- |
| Baseline predictor | n | % | Odds ratio | 95% CI | P-value |
| Age: |  |  |  |  | <0.001 |
| ≤30 years | 381 | 48.4% | 1 |  |  |
| 31-40 years | 466 | 42.9% | 0.80 | 0.67 to 0.97 | 0.021 |
| 41-50 years | 643 | 39.7% | 0.70 | 0.59 to 0.83 | <0.001 |
| 51-60 years | 716 | 43.8% | 0.83 | 0.70 to 0.98 | 0.032 |
| 61+ years | 180 | 39.7% | 0.70 | 0.55 to 0.89 | 0.003 |
| Gender: |  |  |  |  |  |
| Female | 1960 | 42.9% | 1 |  |  |
| Male | 426 | 42.1% | 0.99 | 0.86 to 1.14 | 0.873 |
| Ethnicity: |  |  |  |  | 0.360 |
| White | 2196 | 42.8% | 1 |  |  |
| Black/African/Caribbean | 55 | 46.6% | 1.16 | 0.81 to 1.68 | 0.421 |
| Asian | 71 | 38.4% | 0.81 | 0.60 to 1.10 | 0.171 |
| Mixed/multiple/other | 64 | 46.4% | 1.14 | 0.81 to 1.61 | 0.447 |
| Clinical role: |  |  |  |  | <0.001 |
| Non-clinical | 873 | 39.1% | 1 |  |  |
| Doctor | 153 | 42.3% | 1.17 | 0.93 to 1.47 | 0.187 |
| Nurse/midwife | 724 | 49.7% | 1.53 | 1.34 to 1.75 | <0.001 |
| Other clinical | 634 | 41.7% | 1.10 | 0.96 to 1.25 | 0.187 |
| COVID risk group: |  |  |  |  |  |
| No | 1320 | 43.0% | 1 |  |  |
| Yes | 494 | 46.6% | 1.19 | 1.02 to 1.37 | 0.022 |
| Mental health (MH) status: |  |  |  |  |  |
| No MH disorder (GHQ12<4) | 821 | 32.3% | 1 |  |  |
| Probable MH disorder (GHQ12≥4) | 1518 | 52.0% | 2.28 | 2.04 to 2.55 | <0.001 |
| Type of trust staff work for: |  |  |  |  |  |
| Acute | 1287 | 43.9% | 1 |  |  |
| Baseline predictor | n | % | Odds ratio | 95% CI | P-value |
| Mental health | 1099 | 41.5% | 0.92 | 0.83 to 1.03 | 0.154 |
| Redeployed outside usual role: |  |  |  |  |  |
| No | 2097 | 42.6% | 1 |  |  |
| Yes | 272 | 43.7% | 1.03 | 0.87 to 1.22 | 0.756 |
| Felt supported by colleagues: |  |  |  |  | <0.001 |
| Extremely | 757 | 34.9% | 1 |  |  |
| Quite a lot | 886 | 43.2% | 1.41 | 1.25 to 1.60 | <0.001 |
| Moderately | 445 | 53.4% | 2.13 | 1.81 to 2.50 | <0.001 |
| A little | 208 | 60.8% | 2.91 | 2.30 to 3.68 | <0.001 |
| Not at all | 50 | 61.0% | 2.98 | 1.90 to 4.70 | <0.001 |
| Felt supported by manager: |  |  |  |  | <0.001 |
| Extremely | 568 | 31.6% | 1 |  |  |
| Quite a lot | 680 | 38.6% | 1.36 | 1.18 to 1.56 | <0.001 |
| Moderately | 500 | 51.1% | 2.26 | 1.92 to 2.65 | <0.001 |
| A little | 386 | 61.0% | 3.37 | 2.79 to 4.06 | <0.001 |
| Not at all | 207 | 68.8% | 4.75 | 3.65 to 6.19 | <0.001 |

CI=Confidence interval. GHQ12=General health questionnaire. All analyses are adjusted for age, gender, and ethnicity. Numbers and analyses only include patients with complete baseline age, gender and ethnicity data, complete outcome data, and who completed the long questionnaire.

### Non-COVID-19 related sick leave

Older participants had lower odds of taking non-COVID-19 related sick leave than younger participants (p<0.001), but if they took it, it was slightly longer (p<0.001; Table 4). Males had lower odds of taking non-COVID-19 sick leave (OR=0.73, 95%CI 0.65-0.82), but if they took it, their lengths of leave were similar to females (p=0.770). Doctors were less likely to take non-COVID-19 sick leave than non-clinical colleagues (OR=0.64, 95%CI 0.53-0.78), and where they did, it was shorter (GMR=0.76, 95%CI 0.62-0.93). Whereas, nurses/midwives and other clinical staff had higher odds of taking non-COVID-19 sick leave than non-clinical staff (OR=1.32 [95%CI 1.18-1.48] and OR=1.32 [95%CI 1.18-1.47], respectively), and were off sick for longer (Table 4).

**Table 4:** Non-COVID-19 related sick days in previous 12-months.

| Non-COVID sick days | Number with outcome |  | Dichotomised analyses of any sick days vs. none |  |  | Continuous analyses of participants with at least 1 sick day reported |  |  |
| --- | --- | --- | --- | --- | --- | --- | --- | --- |
| Baseline predictor | n | % | OR | 95% CI | P-value | GMR | 95% CI | P-value |
| Age: |  |  |  |  | <0.001 |  |  | <0.001 |
| ≤30 years | 647 | 60.5% | 1 |  |  | 1 |  |  |
| 31-40 years | 868 | 55.4% | 0.83 | 0.71 to 0.97 | 0.020 | 0.98 | 0.86 to 1.11 | 0.747 |
| 41-50 years | 1091 | 47.2% | 0.59 | 0.51 to 0.68 | <0.001 | 1.02 | 0.90 to 1.16 | 0.744 |
| 51-60 years | 1102 | 45.4% | 0.55 | 0.47 to 0.63 | <0.001 | 1.24 | 1.09 to 1.41 | 0.001 |
| 61+ years | 255 | 40.7% | 0.46 | 0.37 to 0.56 | <0.001 | 1.17 | 0.96 to 1.43 | 0.118 |
| Gender: |  |  |  |  |  |  |  |  |
| Female | 3357 | 51.0% | 1 |  |  | 1 |  |  |
| Male | 606 | 42.7% | 0.73 | 0.65 to 0.82 | <0.001 | 0.98 | 0.88 to 1.10 | 0.770 |
| Ethnicity: |  |  |  |  | 0.307 |  |  | 0.656 |
| White | 3576 | 49.4% | 1 |  |  | 1 |  |  |
| Black/African/Caribbean | 99 | 45.8% | 0.92 | 0.70 to 1.22 | 0.568 | 0.92 | 0.71 to 1.19 | 0.511 |
| Asian | 166 | 51.6% | 1.03 | 0.82 to 1.29 | 0.797 | 1.11 | 0.90 to 1.37 | 0.315 |
| Mixed/multiple/other | 122 | 56.2% | 1.29 | 0.98 to 1.69 | 0.073 | 1.07 | 0.84 to 1.35 | 0.594 |
| Clinical role: |  |  |  |  | <0.001 |  |  | <0.001 |
| Non-clinical | 1473 | 45.7% | 1 |  |  | 1 |  |  |
| Doctor | 183 | 34.5% | 0.64 | 0.53 to 0.78 | <0.001 | 0.76 | 0.62 to 0.93 | 0.008 |
| Nurse/midwife | 1063 | 53.4% | 1.32 | 1.18 to 1.48 | <0.001 | 1.20 | 1.08 to 1.34 | 0.001 |
| Other clinical | 1242 | 55.1% | 1.32 | 1.18 to 1.47 | <0.001 | 1.11 | 1.00 to 1.23 | 0.048 |
| COVID risk group: |  |  |  |  |  |  |  |  |
| No | 1884 | 48.2% | 1 |  |  | 1 |  |  |
| Yes | 769 | 54.4% | 1.46 | 1.29 to 1.67 | <0.001 | 1.43 | 1.27 to 1.62 | <0.001 |
| Mental health (MH) status: |  |  |  |  |  |  |  |  |
| No MH disorder (GHQ12<4) | 1576 | 43.4% | 1 |  |  | 1 |  |  |
| Probable MH disorder (GHQ12≥4) | 2293 | 54.9% | 1.54 | 1.41 to 1.69 | <0.001 | 1.39 | 1.28 to 1.52 | <0.001 |
| Type of trust staff work for: |  |  |  |  |  |  |  |  |
| Acute | 2008 | 49.5% | 1 |  |  | 1 |  |  |
| Baseline predictor | n | % | OR | 95% CI | P-value | GMR | 95% CI | P-value |
| Mental health | 1955 | 49.6% | 0.97 | 0.88 to 1.06 | 0.446 | 1.14 | 1.05 to 1.25 | 0.003 |
| Redeployed outside usual role: |  |  |  |  |  |  |  |  |
| No | 3483 | 49.3% | 1 |  |  | 1 |  |  |
| Yes | 455 | 51.5% | 1.07 | 0.93 to 1.23 | 0.36 | 1.09 | 0.95 to 1.25 | 0.199 |
| Felt supported by colleagues: |  |  |  |  | <0.001 |  |  | <0.001 |
| Extremely | 1438 | 47.1% | 1 |  |  | 1 |  |  |
| Quite a lot | 1459 | 49.2% | 1.07 | 0.96 to 1.18 | 0.205 | 0.95 | 0.86 to 1.05 | 0.318 |
| Moderately | 654 | 53.4% | 1.25 | 1.09 to 1.43 | 0.001 | 1.20 | 1.05 to 1.37 | 0.006 |
| A little | 273 | 55.7% | 1.39 | 1.15 to 1.69 | 0.001 | 1.44 | 1.19 to 1.74 | <0.001 |
| Not at all | 64 | 58.7% | 1.62 | 1.09 to 2.41 | 0.018 | 1.8 | 1.27 to 2.56 | 0.001 |
| Felt supported by manager: |  |  |  |  | <0.001 |  |  | <0.001 |
| Extremely | 1195 | 46.8% | 1 |  |  | 1 |  |  |
| Quite a lot | 1247 | 48.0% | 1.04 | 0.93 to 1.16 | 0.474 | 0.97 | 0.87 to 1.08 | 0.592 |
| Moderately | 737 | 52.1% | 1.21 | 1.06 to 1.38 | 0.004 | 1.10 | 0.97 to 1.25 | 0.156 |
| A little | 493 | 56.7% | 1.46 | 1.25 to 1.71 | <0.001 | 1.14 | 0.98 to 1.31 | 0.089 |
| Not at all | 210 | 53.0% | 1.28 | 1.03 to 1.59 | 0.024 | 1.76 | 1.44 to 2.16 | <0.001 |

OR=Odds ratio. GMR=Geometric mean ratio. CI=Confidence interval. GHQ12=General health questionnaire. All analyses are adjusted for age, gender, and ethnicity. Numbers and analyses only include patients with complete baseline age, gender and ethnicity data, and complete outcome data.

Participants in COVID-19 risk groups had higher odds of taking non-COVID sick leave (OR=1.46, 95%CI 1.29-1.67) and if they did, it was also longer (GMR=1.43, 95%CI 1.27-1.62). Similarly, participants with a probable mental health disorder had higher odds of taking sick leave and for longer periods than participants with no mental health disorder (OR=1.54 [95%CI 1.41-1.69] and GMR=1.39 [95%CI 1.28-1.52], respectively). Participants working in mental health Trusts had similar odds of taking non-COVID-19 sick leave as those in acute Trusts (p=0.446), but if they did take it, it was slightly longer (GMR=1.14, 95%CI 1.05-1.25). Participants with less colleague (p<0.001) and manager (p<0.001) support tended to have higher odds of taking sick leave, and longer periods of sick leave (p<0.001), than participants with better support (Table 4).

### COVID-19 related sick leave

Older participants had lower odds of taking COVID-19 related sick leave than younger participants (p<0.001), but where they did, it was for longer periods (p<0.001; Table 5). All non-white participants had higher odds of COVID-19 sick leave than white participants (p<0.001), with participants of Asian ethnicity having the highest odds (OR=1.63, 95%CI 1.28-2.08). Asian participants also had longer periods of COVID-19 sick leave than white participants (GMR=1.37, 95%CI 1.11-1.69). Doctors, nurses/midwives and other clinical staff all had higher odds of taking COVID-19 related sick leave than non-clinical staff (p<0.001), and nurses/midwives and other clinical staff also had longer periods of COVID-19 related absence (p<0.001; Table 5).

**Table 5:**
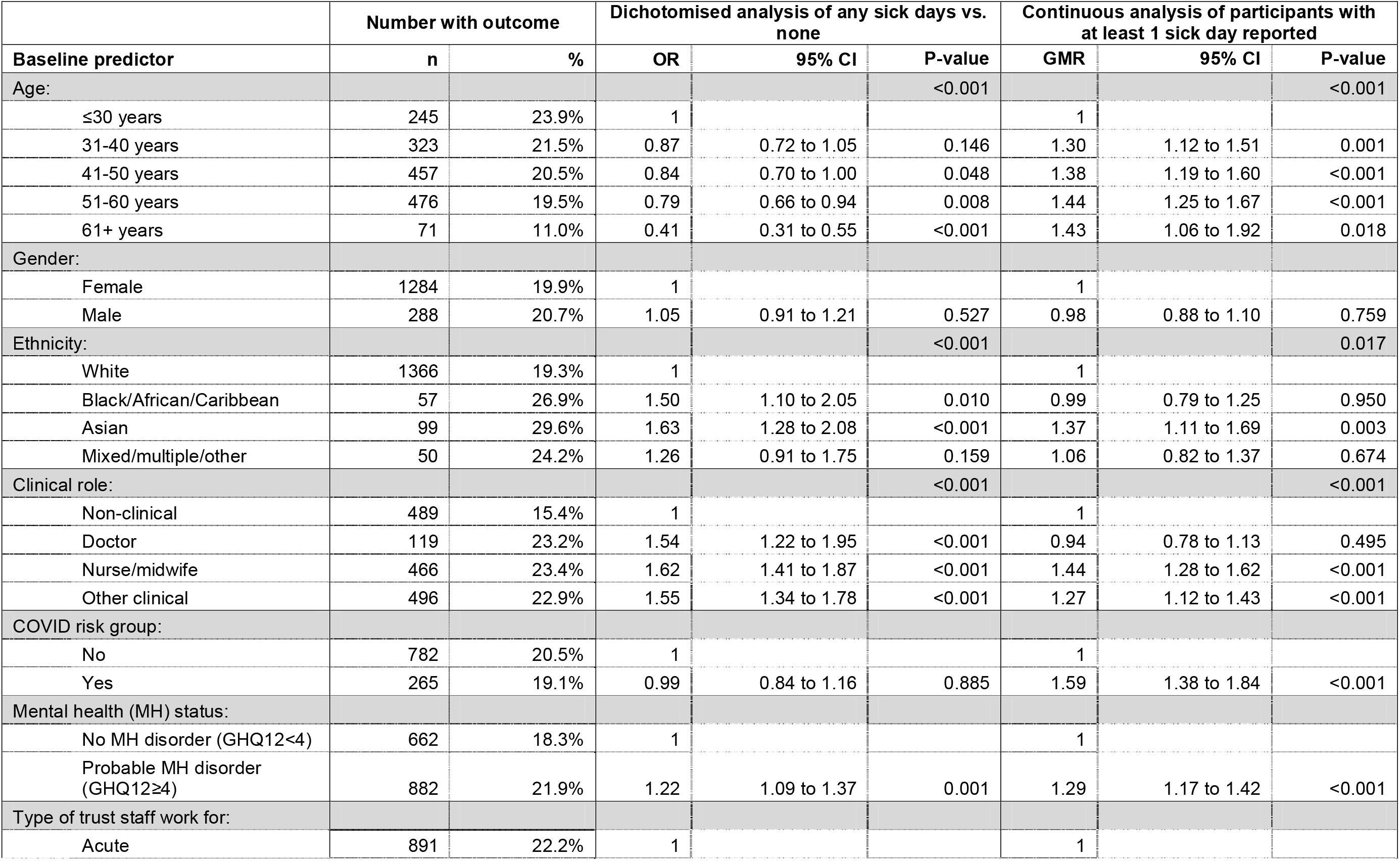
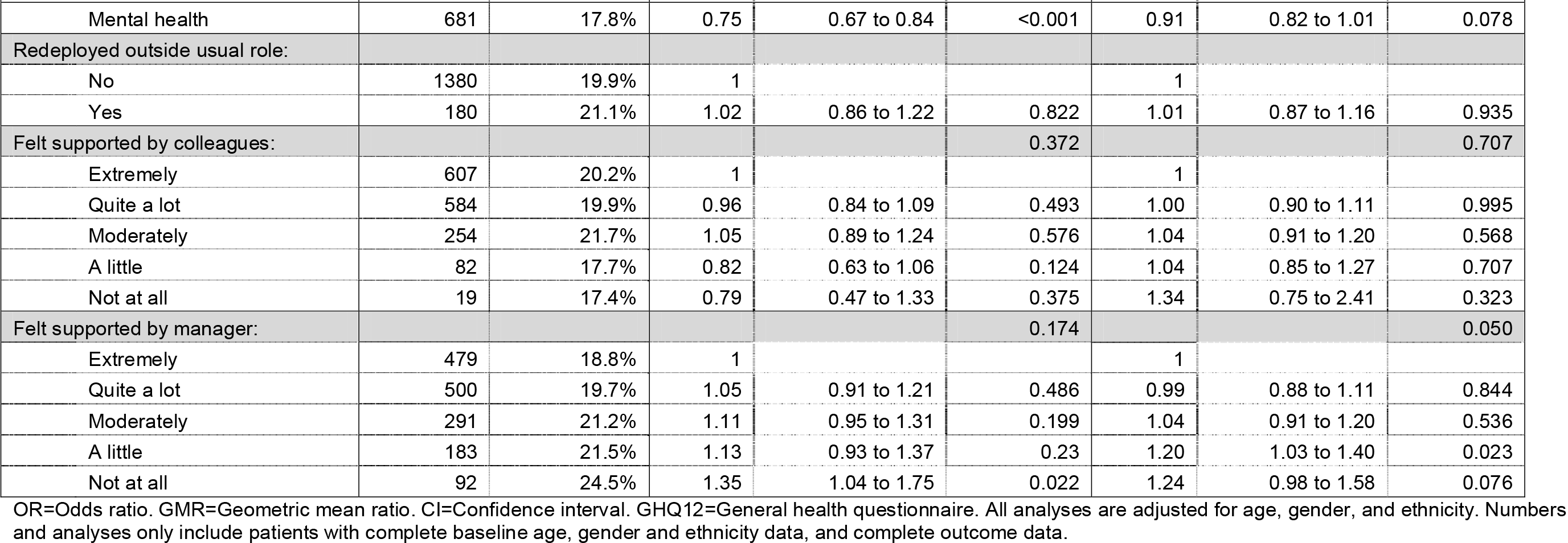
COVID-19 related sick days.

The odds of COVID-19 related sick leave was not associated with COVID-19 risk group (p=0.885), but if they did take it, those in a high-risk group took longer leave (GMR=1.59, 95%CI 1.38-1.84). Participants with a probable mental health disorder had higher odds of taking COVID-19 related sick leave (OR=1.22, 95%CI 1.09-1.37) and had longer leave if they did take it (GMR=1.29, 95%CI 1.17-1.42). Participants working at a mental health Trust had lower odds of taking COVID-19 related sick leave than those at an acute Trust (OR=0.75, 95%CI 0.67-0.84), and slightly shorter periods of leave if they did take it (GMR=0.91, 95%CI 0.82-1.01). Manager support was not associated with taking COVID-19 related leave, but if it was taken, those who felt less supported took slightly longer leave (p=0.050; Table 5).

## Discussion

### Summary of findings

This paper aimed to identify factors associated with staff intention to leave their NHS role and sickness absence. The profile of characteristics associated with these two outcomes were very similar; outcomes were more likely among NHS staff who were younger, in a COVID-19 risk group, had a probable mental health disorder, and who did not feel supported by colleagues and managers.

The association with age may be explained by ‘healthy worker survivor bias’, i.e. healthier workers, and those willing and able to cope with the pressures inherent in the role, remain working, while those who become unwell take more sickness absences and/or leave the workforce (14). Our findings about those in COVID-19 high risk groups are in line with the fact that the majority of high risk participants had a pre-existing health condition (e.g. cardiovascular disease, diabetes, or immune system diseases) (15), making it more likely for them to take sickness absence, and to consider leaving high-risk roles (16). There is a large body of evidence that those with poor mental health are more likely to take sickness absence, and to exit the workforce (7, 8), which is in keeping with our findings here. Our findings about those who feel unsupported by colleagues and managers, are in line with other evidence that poorer perceived leadership and peer support are associated with both higher intention to leave (17) and sickness absence (18).

There were some differences in characteristics between those thinking about or actively intending to leave. Nurses/midwives were more likely than non-clinical staff to be thinking about leaving, while doctors (compared to non-clinical staff) and male participants were less likely to be actively seeking other roles. Given their long training, doctors may be less likely to seek other employment than non-clinical staff, who may perceive wider employment opportunities outside healthcare. The pressures inherent in nursing roles may be the reason they are more likely than non-clinical staff to consider leaving. The COVID-19 pandemic may have had a greater impact on women leaving the paid workforce due to their greater contribution to unpaid care work (19), which may explain our finding that male staff were less likely than female staff to be seeking a new role.

Similarly, there were some differences in characteristics between those taking sickness absence for non-COVID-19 and COVID-19 reasons. Female staff were more likely to take non-COVID-19 sickness absence, as were nurses/midwifes or other clinical staff compared to non-clinical staff. Sickness absence rates have been historically higher for women (20), which is echoed in our findings. The fact that nurses/midwives and other clinical staff were more likely to take sickness absence may be due to the ability of some non-clinical staff to work from home, even when feeling unwell, whereas this is less feasible in clinical roles.

In terms of COVID-19 sickness absence, staff of non-white ethnicities were more likely to be absent, as were those in clinical roles (compared to non-clinical roles), and those working in an acute hospital rather than mental health settings. There is evidence of higher risk of infection with COVID-19 and of worse clinical outcomes for people from minoritized ethnic backgrounds compared to white people (21), which is in line with our findings. Similar reasoning may apply as non-COVID-19 absence regarding non-clinical staff being more able to work from home while isolating or unwell with COVID-19 than clinical staff. Similarly, acute hospital workers were more likely to be on site facing higher infection risks (so required more COVID-19 sickness absence) than those in mental health settings who were more likely to be able to move services to virtual settings.

### Strengths and limitations of the study

The main strength of this study is the sample size, with participants from 18 acute and mental health NHS Trusts across England, in differing areas of deprivation, with a mixture of urban and rural settings, and a response rate of 15%. Another strength is the longitudinal nature of the data analysed. The availability of follow-up data has enabled us to explore predictive factors in a robust way, which is not possible with cross-sectional data used by most research in this area. We present data on both intention to leave, and sickness absence, two major factors involved in the current NHS workforce crisis (4).

However, there is some potential bias in our sample. Around half of the participants (11,724/22,555; 52%) who completed the baseline questionnaire did not complete the 12-month follow-up questionnaire, and there was substantial missingness within the second part of the 12-month questionnaire (which included the ‘intention to leave’ outcome questions). We present the baseline characteristics for each of the cohorts of data and modelled which exposures were associated with missingness in each of the outcomes. Findings were generally in the same direction as the effects in the outcome models, which, if anything, is likely to have led to an underestimation of associations between exposures and outcomes rather than an overestimation.

This study was carried out during the COVID-19 pandemic and therefore, given the heightened staff stress during that time and the large turnover of staff since then, the proportions of people taking sick leave and intending to leave the NHS may differ from now; however, we believe our findings are still likely to be generalisable to the NHS workforce now. Unfortunately, we do not have pre-pandemic data for this cohort, so are unable to provide insight to any changing trends from before this data was collected.

### Implications for research and/or practice

Further research in this area could usefully explore methodological advances aimed at accounting for healthy worker survivor bias (14, 22). The ability to link cohort data to pre-pandemic health and wellbeing, sickness absence, and retention data should be explored, as this could offer insights to the long-term trajectories of these outcomes.

The clear stepwise association between how well supported someone feels by their colleagues and managers and likelihood of actively or thinking about job seeking outside their profession has important implications for employers. There are likely to be considerable benefits from training managers to speak with and support staff, especially those who are experiencing mental health difficulties. Furthermore, ensuring teams have sufficient opportunities to form and foster social connections and reflect on the challenges of their work together, may reduce the likelihood of staff leaving their role or taking excessive sick days. This is already partially addressed in the recently published NHS long term workforce plan (9).

## Data Availability

Data will be available to researchers who provide a justified hypothesis and structured statistical analysis plan addressing a legitimate research question that is approved by the NHS CHECK Senior Research Team and after the signing of a data sharing agreement. Only deidentified participant data will be provided.

## Acknowledgements

We are especially grateful to all the participants who took part in the study. We wish to acknowledge the National Institute of Health Research (NIHR) Applied Research Collaboration (ARC) National NHS and Social Care Workforce Group, with the following ARCs: East Midlands, East of England, South-West Peninsula, South London, West, North-West Coast, Yorkshire and Humber, and North-East and North Cumbria. They enabled the set-up of the national network of participating hospital sites and aided the research team to recruit effectively during the COVID-19 pandemic.

The NHS CHECK consortium includes the following site leads: Siobhan Coleman, Sean Cross, Amy Dewar, Chris Dickens, Frances Farnworth, Adam Gordon, Charles Goss, Jessica Harvey, Nusrat Husain, Peter Jones, Damien Longson, Paul Moran, Jesus Perez, Mark Pietroni, Ian Smith, Tayyeb Tahir, Peter Trigwell, Jeremy Turner, Julian Walker, Scott Weich, Ashley Wilkie.

The NHS CHECK consortium includes the following co-investigators and collaborators: Peter Aitken, Ewan Carr, Anthony David, Mary Jane Doherty, Sarah Dorrington, Rosie Duncan, Sam Gnanapragasam, Cerisse Gunasinghe, Stephani Hatch, Danielle Lamb, Daniel Leightley, Ira Madan, Richard Morriss, Isabel McMullen, Dominic Murphy, Martin Parsons, Catherine Polling, Alexandra Pollitt, Anne-Marie Rafferty, Rebecca Rhead, Danai Serfioti, Chloe Simela, Charlotte Wilson Jones.

