## Supplementary Table for "NHS staff: Sickness absence and intention to leave the profession"

**Supplementary material**

**Table S1 Participant baseline characteristics for participants who reported data for each of the four outcomes**

|  | | **Whole cohort**  **(n=22,438)** | | **12-month cohort**  **(n=10,776)** | | **Non-COVID sick leave (n=8,320)** | | **COVID sick leave**  **(n=8,198)** | | **Actively seeking new role (n=5,795)** | | **Thinking about leaving (n=5,805)** | |
| --- | --- | --- | --- | --- | --- | --- | --- | --- | --- | --- | --- | --- | --- |
|  | | **n** | **%** | **n** | **%** | **n** | **%** | **n** | **%** | **n** | **%** | **n** | **%** |
| **Age (median, IQR)** | | 43 | (33, 53) | 47 | (36, 54) | 47 | (37, 54) | 47 | (37, 55) | 47 | (36, 54) | 47 | (36, 54) |
|  | ≤30 years | 4273 | 19.0% | 1485 | 13.8% | 1070 | 12.9% | 1023 | 12.5% | 788 | 13.6% | 787 | 13.6% |
|  | 31-40 years | 4915 | 21.9% | 2019 | 18.7% | 1567 | 18.8% | 1503 | 18.3% | 1087 | 18.8% | 1087 | 18.7% |
|  | 41-50 years | 5620 | 25.0% | 2904 | 26.9% | 2311 | 27.8% | 2231 | 27.2% | 1617 | 27.9% | 1618 | 27.9% |
|  | 51-60 years | 5272 | 23.5% | 3097 | 28.7% | 2433 | 29.2% | 2450 | 29.9% | 1633 | 28.2% | 1638 | 28.2% |
|  | ≥61 years | 1328 | 5.9% | 802 | 7.4% | 626 | 7.5% | 646 | 7.9% | 449 | 7.7% | 453 | 7.8% |
|  | Missing | 1030 | 4.6% | 469 | 4.4% | 313 | 3.8% | 345 | 4.2% | 221 | 3.8% | 222 | 3.8% |
| **Gender** | |  |  |  |  |  |  |  |  |  |  |  |  |
|  | Female | 18125 | 80.8% | 8785 | 81.5% | 6839 | 82.2% | 6744 | 82.3% | 4737 | 81.7% | 4747 | 81.8% |
|  | Male | 4177 | 18.6% | 1964 | 18.2% | 1462 | 17.6% | 1437 | 17.5% | 1043 | 18.0% | 1043 | 18.0% |
|  | Missing | 136 | 0.6% | 27 | 0.3% | 19 | 0.2% | 17 | 0.2% | 15 | 0.3% | 15 | 0.3% |
| **Ethnicity** | |  |  |  |  |  |  |  |  |  |  |  |  |
|  | White | 19093 | 85.1% | 9604 | 89.1% | 7506 | 90.2% | 7381 | 90.0% | 5315 | 91.7% | 5326 | 91.7% |
|  | Black | 969 | 4.3% | 334 | 3.1% | 228 | 2.7% | 225 | 2.7% | 124 | 2.1% | 124 | 2.1% |
|  | Asian | 1465 | 6.5% | 484 | 4.5% | 334 | 4.0% | 351 | 4.3% | 190 | 3.3% | 190 | 3.3% |
|  | Mixed | 539 | 2.4% | 238 | 2.2% | 173 | 2.1% | 167 | 2.0% | 104 | 1.8% | 103 | 1.8% |
|  | Other | 199 | 0.9% | 72 | 0.7% | 51 | 0.6% | 48 | 0.6% | 38 | 0.7% | 38 | 0.7% |
|  | Missing | 173 | 0.8% | 44 | 0.4% | 28 | 0.3% | 26 | 0.3% | 24 | 0.4% | 24 | 0.4% |
| **Clinical role** | |  |  |  |  |  |  |  |  |  |  |  |  |
|  | Doctor | 1623 | 7.2% | 704 | 6.5% | 548 | 6.6% | 531 | 6.5% | 373 | 6.4% | 373 | 6.4% |
|  | Nurse/midwife | 5707 | 25.4% | 2717 | 25.2% | 2061 | 24.8% | 2077 | 25.3% | 1500 | 25.9% | 1502 | 25.9% |
|  | Other clinical | 6601 | 29.4% | 2983 | 27.7% | 2331 | 28.0% | 2256 | 27.5% | 1572 | 27.1% | 1576 | 27.1% |
|  | Non-clinical | 8384 | 37.4% | 4345 | 40.3% | 3360 | 40.4% | 3318 | 40.5% | 2335 | 40.3% | 2339 | 40.3% |
|  | Missing | 123 | 0.5% | 27 | 0.3% | 20 | 0.2% | 16 | 0.2% | 15 | 0.3% | 15 | 0.3% |
| **Covid risk group** | |  |  |  |  |  |  |  |  |  |  |  |  |
|  | No | 9124 | 40.7% | 5082 | 47.2% | 4052 | 48.7% | 3969 | 48.4% | 3178 | 54.8% | 3182 | 54.8% |
|  | Yes | 3161 | 14.1% | 1875 | 17.4% | 1473 | 17.7% | 1454 | 17.7% | 1104 | 19.1% | 1104 | 19.0% |
|  | Missing | 10153 | 45.2% | 3819 | 35.4% | 2795 | 33.6% | 2775 | 33.8% | 1513 | 26.1% | 1519 | 26.2% |
| **Mental health (MH) status** | |  |  |  |  |  |  |  |  |  |  |  |  |
|  | No MH disorder (GHQ12<4) | 10075 | 44.9% | 4903 | 45.5% | 3781 | 45.4% | 3790 | 46.2% | 2643 | 45.6% | 2651 | 45.7% |
|  | Probable MH disorder (GHQ12≥4) | 11254 | 50.2% | 5529 | 51.3% | 4311 | 51.8% | 4184 | 51.0% | 3018 | 52.1% | 3020 | 52.0% |
|  | Missing | 1109 | 4.9% | 344 | 3.2% | 228 | 2.7% | 224 | 2.7% | 134 | 2.3% | 134 | 2.3% |
| **Trust type** | |  |  |  |  |  |  |  |  |  |  |  |  |
|  | Acute trust | 11241 | 50.1% | 5549 | 51.5% | 4244 | 51.0% | 4228 | 51.6% | 3065 | 52.9% | 3073 | 52.9% |
|  | Mental Health trust | 11197 | 49.9% | 5227 | 48.5% | 4076 | 49.0% | 3970 | 48.4% | 2730 | 47.1% | 2732 | 47.1% |
| **Redeployed outside usual role** | |  |  |  |  |  |  |  |  |  |  |  |  |
|  | No | 19340 | 86.2% | 9468 | 87.9% | 7345 | 88.3% | 7248 | 88.4% | 5116 | 88.3% | 5125 | 88.3% |
|  | Yes | 2765 | 12.3% | 1207 | 11.2% | 910 | 10.9% | 886 | 10.8% | 637 | 11.0% | 637 | 11.0% |
|  | Missing | 333 | 1.5% | 101 | 0.9% | 65 | 0.8% | 64 | 0.8% | 42 | 0.7% | 43 | 0.7% |
| **Supported by colleagues** | |  |  |  |  |  |  |  |  |  |  |  |  |
|  | Extremely | 8334 | 37.1% | 4086 | 37.9% | 3179 | 38.2% | 3159 | 38.5% | 2258 | 39.0% | 2267 | 39.1% |
|  | Quite a bit | 8189 | 36.5% | 3984 | 37.0% | 3075 | 37.0% | 3045 | 37.1% | 2120 | 36.6% | 2120 | 36.5% |
|  | Moderately | 3420 | 15.2% | 1618 | 15.0% | 1258 | 15.1% | 1209 | 14.7% | 859 | 14.8% | 860 | 14.8% |
|  | A little bit | 1367 | 6.1% | 657 | 6.1% | 507 | 6.1% | 485 | 5.9% | 353 | 6.1% | 353 | 6.1% |
|  | Not at all | 311 | 1.4% | 157 | 1.5% | 116 | 1.4% | 115 | 1.4% | 90 | 1.6% | 89 | 1.5% |
|  | Missing | 817 | 3.6% | 274 | 2.5% | 185 | 2.2% | 185 | 2.3% | 115 | 2.0% | 116 | 2.0% |
| **Supported by manager** | |  |  |  |  |  |  |  |  |  |  |  |  |
|  | Extremely | 7052 | 31.4% | 3443 | 32.0% | 2670 | 32.1% | 2668 | 32.5% | 1872 | 32.3% | 1878 | 32.4% |
|  | Quite a bit | 7040 | 31.4% | 3409 | 31.6% | 2681 | 32.2% | 2635 | 32.1% | 1814 | 31.3% | 1816 | 31.3% |
|  | Moderately | 3989 | 17.8% | 1912 | 17.7% | 1467 | 17.6% | 1424 | 17.4% | 1015 | 17.5% | 1017 | 17.5% |
|  | A little bit | 2374 | 10.6% | 1164 | 10.8% | 897 | 10.8% | 883 | 10.8% | 653 | 11.3% | 653 | 11.2% |
|  | Not at all | 1141 | 5.1% | 563 | 5.2% | 411 | 4.9% | 395 | 4.8% | 318 | 5.5% | 317 | 5.5% |
|  | Missing | 842 | 3.8% | 285 | 2.6% | 194 | 2.3% | 193 | 2.4% | 123 | 2.1% | 124 | 2.1% |

IQR=Interquartile range. GHQ12=General health questionnaire. Note. Participants who reported their gender as other or prefer not to say have been excluded from this table as numbers were too small to include in analysis models.

**Table S2 Predicting missingness of job hunt and sick leave questions in the 12-month questionnaire**

|  | | Actively seeking a new job | | | Regularly thinking about leaving profession | | | Non-COVID-19 sick leave | | | COVID-19 sick leave | | |
| --- | --- | --- | --- | --- | --- | --- | --- | --- | --- | --- | --- | --- | --- |
| Baseline predictor | | OR | 95% CI | P-value | OR | 95% CI | P-value | OR | 95% CI | P-value | OR | 95% CI | P-value |
| Age: | |  |  | <0.001 |  |  | <0.001 |  |  | <0.001 |  |  | <0.001 |
|  | ≤30 years | 1 |  |  | 1 |  |  | 1 |  |  | 1 |  |  |
|  | 31-40 years | 0.81 | 0.73 to 0.90 | <0.001 | 0.81 | 0.73 to 0.90 | <0.001 | 0.72 | 0.66 to 0.79 | <0.001 | 0.72 | 0.66 to 0.79 | <0.001 |
|  | 41-50 years | 0.57 | 0.52 to 0.63 | <0.001 | 0.57 | 0.52 to 0.63 | <0.001 | 0.49 | 0.44 to 0.53 | <0.001 | 0.48 | 0.44 to 0.53 | <0.001 |
|  | 51-60 years | 0.54 | 0.49 to 0.59 | <0.001 | 0.53 | 0.48 to 0.59 | <0.001 | 0.41 | 0.37 to 0.45 | <0.001 | 0.38 | 0.34 to 0.41 | <0.001 |
|  | 61+ years | 0.47 | 0.41 to 0.54 | <0.001 | 0.47 | 0.41 to 0.54 | <0.001 | 0.39 | 0.35 to 0.45 | <0.001 | 0.35 | 0.30 to 0.39 | <0.001 |
| Gender: | |  |  |  |  |  |  |  |  |  |  |  |  |
|  | Female | 1 |  |  | 1 |  |  | 1 |  |  | 1 |  |  |
|  | Male | 1.03 | 0.96 to 1.12 | 0.407 | 1.04 | 0.96 to 1.12 | 0.367 | 1.12 | 1.04 to 1.20 | 0.003 | 1.13 | 1.05 to 1.21 | 0.001 |
| Ethnicity: | |  |  | <0.001 |  |  | <0.001 |  |  | <0.001 |  |  | <0.001 |
|  | White | 1 |  |  | 1 |  |  | 1 |  |  | 1 |  |  |
|  | Black/African/Caribbean | 2.60 | 2.13 to 3.16 | <0.001 | 2.60 | 2.14 to 3.17 | <0.001 | 2.09 | 1.78 to 2.44 | <0.001 | 2.06 | 1.76 to 2.41 | <0.001 |
|  | Asian | 2.25 | 1.91 to 2.63 | <0.001 | 2.25 | 1.92 to 2.64 | <0.001 | 1.84 | 1.61 to 2.09 | <0.001 | 1.66 | 1.46 to 1.89 | <0.001 |
|  | Mixed/multiple/other | 1.50 | 1.24 to 1.81 | <0.001 | 1.51 | 1.25 to 1.83 | <0.001 | 1.36 | 1.15 to 1.60 | <0.001 | 1.39 | 1.17 to 1.64 | <0.001 |
| Clinical role: | |  |  | 0.011 |  |  | 0.013 |  |  | 0.014 |  |  | 0.015 |
|  | Non-clinical | 1 |  |  | 1 |  |  | 1 |  |  | 1 |  |  |
|  | Doctor | 1.10 | 0.96 to 1.25 | 0.177 | 1.10 | 0.96 to 1.25 | 0.173 | 1.10 | 0.98 to 1.24 | 0.105 | 1.13 | 1.01 to 1.28 | 0.039 |
|  | Nurse/midwife | 1.01 | 0.93 to 1.09 | 0.871 | 1.01 | 0.93 to 1.09 | 0.852 | 1.11 | 1.04 to 1.20 | 0.004 | 1.07 | 1.00 to 1.15 | 0.062 |
|  | Other clinical | 1.13 | 1.04 to 1.22 | 0.003 | 1.12 | 1.04 to 1.22 | 0.003 | 1.09 | 1.02 to 1.17 | 0.018 | 1.11 | 1.03 to 1.19 | 0.004 |
| COVID risk group: | |  |  |  |  |  |  |  |  |  |  |  |  |
|  | No | 1 |  |  | 1 |  |  | 1 |  |  | 1 |  |  |
|  | Yes | 1.06 | 0.97 to 1.16 | 0.214 | 1.06 | 0.97 to 1.16 | 0.190 | 1.00 | 0.92 to 1.09 | 0.981 | 1.01 | 0.93 to 1.11 | 0.780 |
| Mental health (MH) status: | |  |  |  |  |  |  |  |  |  |  |  |  |
|  | No MH disorder (GHQ12<4) | 1 |  |  | 1 |  |  | 1 |  |  | 1 |  |  |
|  | Probable MH disorder (GHQ12≥4) | 0.95 | 0.89 to 1.01 | 0.123 | 0.96 | 0.90 to 1.02 | 0.152 | 0.94 | 0.88 to 0.99 | 0.024 | 0.98 | 0.92 to 1.04 | 0.449 |
| Type of trust staff work for: | |  |  |  |  |  |  |  |  |  |  |  |  |
|  | Acute | 1 |  |  | 1 |  |  | 1 |  |  | 1 |  |  |
|  | Mental health | 1.29 | 1.22 to 1.38 | <0.001 | 1.30 | 1.22 to 1.38 | <0.001 | 1.20 | 1.13 to 1.27 | <0.001 | 1.23 | 1.16 to 1.30 | <0.001 |
| Redeployed outside usual role: | |  |  |  |  |  |  |  |  |  |  |  |  |
|  | No | 1 |  |  | 1 |  |  | 1 |  |  | 1 |  |  |
|  | Yes | 1.10 | 0.99 to 1.21 | 0.065 | 1.10 | 1.00 to 1.21 | 0.060 | 1.13 | 1.04 to 1.24 | 0.005 | 1.15 | 1.05 to 1.26 | 0.002 |
| Felt supported by colleagues: | |  |  | 0.601 |  |  | 0.602 |  |  | 0.997 |  |  | 0.618 |
|  | Extremely | 1 |  |  | 1 |  |  | 1 |  |  | 1 |  |  |
|  | Quite a lot | 1.03 | 0.96 to 1.11 | 0.392 | 1.04 | 0.97 to 1.12 | 0.303 | 1 | 0.93 to 1.06 | 0.875 | 0.99 | 0.93 to 1.06 | 0.747 |
|  | Moderately | 1.05 | 0.95 to 1.15 | 0.353 | 1.05 | 0.96 to 1.15 | 0.311 | 0.99 | 0.91 to 1.08 | 0.808 | 1.05 | 0.96 to 1.14 | 0.309 |
|  | A little | 1.01 | 0.88 to 1.15 | 0.927 | 1.01 | 0.89 to 1.16 | 0.863 | 0.98 | 0.87 to 1.11 | 0.800 | 1.06 | 0.93 to 1.20 | 0.377 |
|  | Not at all | 0.87 | 0.67 to 1.13 | 0.279 | 0.89 | 0.68 to 1.15 | 0.364 | 0.97 | 0.75 to 1.24 | 0.777 | 0.94 | 0.74 to 1.21 | 0.638 |
| Felt supported by manager: | |  |  | 0.120 |  |  | 0.140 |  |  | 0.534 |  |  | 0.259 |
|  | Extremely | 1 |  |  | 1 |  |  | 1 |  |  | 1 |  |  |
|  | Quite a lot | 1.02 | 0.94 to 1.10 | 0.636 | 1.02 | 0.95 to 1.10 | 0.584 | 0.96 | 0.89 to 1.03 | 0.243 | 0.99 | 0.92 to 1.06 | 0.693 |
|  | Moderately | 1.03 | 0.94 to 1.13 | 0.523 | 1.03 | 0.94 to 1.13 | 0.479 | 1.01 | 0.93 to 1.10 | 0.769 | 1.05 | 0.97 to 1.15 | 0.209 |
|  | A little | 0.91 | 0.82 to 1.01 | 0.082 | 0.91 | 0.82 to 1.02 | 0.097 | 0.95 | 0.86 to 1.05 | 0.312 | 0.97 | 0.88 to 1.07 | 0.563 |
|  | Not at all | 0.90 | 0.78 to 1.04 | 0.164 | 0.91 | 0.79 to 1.05 | 0.205 | 1.02 | 0.89 to 1.17 | 0.747 | 1.10 | 0.96 to 1.27 | 0.159 |

OR=Odds ratio. CI=Confidence interval. GHQ12=General health questionnaire.
